# microRNA-based predictor for diagnosis of frontotemporal dementia

**DOI:** 10.1101/2020.01.22.20018408

**Authors:** Iddo Magen, Nancy-Sarah Yacovzada, Jason D. Warren, Carolin Heller, Imogen Swift, Yoana Bobeva, Andrea Malaspina, Jonathan D. Rohrer, Pietro Fratta, Eran Hornstein

**Affiliations:** Department of Molecular Genetics, Weizmann Institute of Science, Rehovot, Israel; Department of Molecular Neuroscience, Weizmann Institute of Science, Rehovot, Israel; Dementia Research Centre, and 4 UK Dementia Research Institute, Department of Neurodegenerative Disease, UCL Queen Square Institute of Neurology, London, UK; Centre for Neuroscience and Trauma, Blizard Institute, Barts and the London School of Medicine and Dentistry, Queen Mary University of London, London, UK; Department of Neuromuscular Diseases, UCL Queen Square Institute of Neurology, London, UK

**Keywords:** frontotemporal dementia, biomarker, microRNA, predictor, feature elimination.

## Abstract

**BACKGROUND:** Frontotemporal dementia (FTD) is an early onset dementia that is diagnosed in ∼20% of the progressive dementia cases. Heterogeneity in FTD clinical presentation too often delays clinical diagnosis and calls for molecular biomarkers to assist diagnosis, including cell free microRNAs (miRNA). However, nonlinearity in the relationship of miRNAs to clinical states and underpowered cohorts has limited research in this domain.

**METHODS:** We initially studied a training cohort of 219 subjects (135 FTD and 84 non-neurodegenerative controls) and then validated the results in a cohort of 74 subjects (33 FTD and 41 controls).

**RESULTS:** Based on cell-free plasma miRNA profiling by next generation sequencing and machine learning approaches, we develop a nonlinear prediction model that accurately distinguishes FTD from non-neurodegenerative controls in ∼90% of cases.

**DISCUSSION:** The fascinating potential of diagnostic miRNA biomarkers might enable early-stage detection and a cost-effective screening approach for clinical trials that can facilitate drug development.

## Background

Frontotemporal dementia (FTD) is a neurodegenerative disease that is characterized by frontal and temporal lobe atrophy, and typically manifests with behavioral or language deficits ^1–4^. The key known genetic drivers of FTD include hexanucleotide repeat expansion in the first intron of the gene chromosome 9 open reading frame 72 (*C9ORF72*) ^5, 6^, and mutations in the genes encoding for microtubule associated protein Tau **(*MAPT*)** ^7^, Valosin-containing protein (*VCP),* TANK-binding kinase 1 *(TBK1),* progranulin (*GRN*), charged multivesicular body protein 2B (*CHMP2B*) and the RNA-binding protein TAR DNA-binding protein 43 (*TDP-43*) ^8–12^. Relatedly, inclusions of Tau and TDP-43 are hallmarks of FTD neuropathology^13, 14^.

FTD can be difficult to diagnose, due to heterogeneity in clinical presentation ^1^. Three main phenotypes of FTD are described: behavioral variant frontotemporal dementia (bvFTD), characterized by changes in social behavior and conduct, semantic dementia (SD), characterized by the loss of semantic knowledge, leading to impaired word comprehension and progressive non-fluent aphasia (PNFA), characterized by progressive difficulties in speech production ^1, 15^. Related motor variants within the FTD spectrum are corticobasal syndrome (CBS) and progressive supranuclear palsy (PSP) ^16^. FTD further resides on a genetic and a clinico-pathological continuum with amyotrophic lateral sclerosis (ALS) ^17^.

Brain imaging and several biofluid proteins have been proposed as biomarkers for FTD ^18–48^. Among the proteins, noticeable are neurofilament light chain (NfL), TDP-43, and phospho-tau, amyloid beta and glial fibrillary acidic protein (GFAP). A recent study concluded that high NfL blood levels are indicative of the intensity of neurodegeneration or the extent of the degenerated axons in FTD ^26^.

microRNAs (miRNAs) are a class of small, non-coding RNAs, that can be quantified in biofluids in a massively parallel fashion, yielding fine-grained profiles ^49^. miRNAs were suggested as biomarkers in neurodegeneration and psychiatry ^50^, and we too have shown their potential as cell-free biomarkers in neurodegeneration, focusing on the motor neuron diseases spinal muscular atrophy (SMA) and ALS ^51, 52^. Thus, low levels of miR-133 and miR-206 in the cerebrospinal fluid (CSF) of patients with SMA, predicted clinically meaningful response to nusinersen therapy ^51^, whereas low plasma levels of miR-181 predict longer survival and slower progression in patients with ALS ^52^.

Several studies suggested plasma or CSF miRNAs as diagnostic biomarkers for FTD^53–62^. However, initial studies of miRNAs in diagnosis of FTD were confounded by cohort size, sample heterogeneity biases or pre-selection of candidate miRNAs. Furthermore, these studies did not address the potential non-linear relationships between miRNAs in developing a predictor.

In the current study, we profiled blood plasma miRNAs, and developed miRNA-based classifier for diagnosing FTD in a training cohort of 219 subjects, that was further validated in another cohort of 74 subjects. We implemented an ensemble machine learning approach, to address biomarker nonlinearity and were able to expose unrevealed disease-associated signals. We then confirmed that these signals are similar between clinical subtypes of FTD. The diagnostic power of the study roots from unbiased miRNA signature discovered by advanced machine learning, on a large and heterogeneous cohort, and validation in an independent held-out cohort, according to the TRIPOD guidelines ^63^. Therefore, circulating miRNAs hold a fascinating potential as diagnostic biomarkers that may shorten diagnostic delay in FTD.

## Methods

### Participants and Sampling

Demographic data of study participants are detailed in Table 1.

**Table 1.** Summary of demographic and clinical characteristics of participants suffering from FTD and control samples. bvFTD: behavioural FTD; PNFA: progressive nonfluent aphasia; SD: semantic dementia. Mean±SD.

|  | Control | FTD |
| --- | --- | --- |
| Number of subjects (% males) | 125 (32%) | 168 (65.5%) |
| UCL controls | 48 (45.8%) |  |
| Queen Mary Hospital controls | 77 (23.4%) |  |
| Age at enrolment | 59.5±10.2 yr. | 65.8±8.1 yr. |
| UCL controls | 65.6±7.3 yr. |  |
| Queen Mary Hospital controls | 55.7±10.0 yr |  |
| Age of onset (1<sup>st</sup> reported symptoms) |  | 60.3±8.3 yr. |
| Disease duration at enrolment |  | 5.5±3.4 yr. |
| FTD clinical subtype<br>(bvFTD/PNFA/SD/FTD-ALS/others) |  | 81/40/28/5/14 |
| FTD Mutation carriers (C9ORF72/MAPT/GRN/TBK1) |  | 18/13/13/2 |
| Likely FTD pathology (TDP-43/Tau) |  | 63/18 |

FTD subjects and their respective controls were enrolled in the longitudinal FTD cohort studies at UCL. Study cohort included 169 FTD patients and 56 controls. Additional controls (N=102) were obtained from the ALS biomarker study (Ethics approval 09/H0703/27) in Queen Mary Hospital, totaling 158 non-neurodegenerative controls. Controls were typically spouses or relatives of patients and were not reported to have any clinical signs of ALS or FTD. Informed consent was obtained from all participants. Study inclusion period was from 2009 to 2018.

All FTD patients were seen at the National Hospital for Neurology and Neurosurgery, a national referral center for young and genetic dementias in the UK; the clinic has high diagnostic accuracy in cases that come to post mortem; for the majority of cases seen in the clinic CSF biomarkers of amyloid and Tau are used to distinguish FTD and AD whenever there is any question over accuracy of diagnosis.

Blood was collected by venipuncture in EDTA tubes, and plasma was recovered from the whole blood sample by centrifugation for 10 minutes at 3500 RPM at 4°C within 1 hour of sampling, and stored at −80°C until RNA extraction and subsequent small RNA next generation sequencing. Frozen plasma samples of FTD and controls from the UCL Biobanks were shipped to the Weizmann Institute of Science for molecular analysis.

### Study Design

Based on power analysis calculations, we found that 150 controls and 150 cases are sufficient to obtain an ROC AUC of 0.7 with a power of 99% and a p-value of 0.0001. Phenotypic data on de-identified patients was separated and blinded during steps of the molecular analysis.

### Small RNA Next Generation Sequencing

Total RNA was extracted from plasma using the miRNeasy micro kit (Qiagen, Hilden, Germany) and quantified with Qubit fluorometer using RNA broad range (BR) assay kit (Thermo Fisher Scientific, Waltham, MA). For small RNA next generation sequencing (RNA-seq), libraries were prepared from 7.5 ng of total RNA using the QIAseq miRNA Library Kit and QIAseq miRNA NGS 48 Index IL (Qiagen), by an experimenter who was blinded to the identity of samples. Samples were randomly allocated to library preparation and sequencing in batches. Precise linear quantification of miRNA is achieved by using unique molecular identifiers (UMIs), of random 12-nucleotide after 3’ and 5’ adapter ligation, within the reverse transcription primers ^49^. cDNA libraries were amplified by PCR for 22 cycles, with a 3’ primer that includes a 6-nucleotide unique index, followed by on-bead size selection and cleaning. Library concentration was determined with Qubit fluorometer (dsDNA high sensitivity assay kit; Thermo Fisher Scientific, Waltham, MA) and library size with Tapestation D1000 (Agilent). Libraries with different indices were multiplexed and sequenced on NextSeq 500/550 v2 flow cell or Novaseq SP100 (Illumina), with 75bp single read and 6bp index read. Fastq files were de-multiplexed using the user-friendly transcriptome analysis pipeline (UTAP) ^64^. Human miRNAs, as defined by miRBase ^65^, were mapped using the GeneGlobe pipeline (https://geneglobe.qiagen.com/us/analyze). We defined "true positive" miRNAs and reduced the likelihood of considering “false positive” miRNAs, following previous works on miRNA biomarkers in neurodegeneration^58^ and other conditions^66–68^. To this end, we included only miRNAs with an average UMI counts > 100 across all samples and with at least a single UMI across all samples, similar to our previous works^51, 52^. Data were further corrected for the library preparation batch in order to reduce its potential bias, and normalized with DESeq2 package ^69^ under the assumption that miRNA counts followed negative binomial distribution

### Constructing Cohorts and Restricting Age and Sex Biases

We observed a younger mean age in controls (53.8±14.5, 95% CI [51.5-56.1]) than in FTD (65.6±8.4, 95% CI [64.4-67.0], Table S1). We reduced age-variance by excluding 34 participants younger than 40, which reduced differences in mean age across the remaining meta-cohort of 293 subjects by 45%. Thus, 168 out of 169 FTD patients and 125 out of 158 non-neurodegenerative control samples were included in the analysis (Table S1). In order to verify that a merged dataset of controls, collected in two different clinical centers, does not introduce biases, we employed the t-distributed stochastic neighbor embedding (t-SNE) algorithm and measured Kullback–Leibler divergence, the difference between probability distributions. In addition, a higher prevalence of males was observed among FTD patients (65%) than among the controls (35%). Therefore, sex and age variables were added to the prediction model as covariates, in addition to the selected 13 miRNA predictors.

### Gradient Boosted Trees for the Development of Disease Binary Classifiers

The FTD-disease binary classifier was developed using Gradient Boosting Classifier, a machine-learning algorithm that uses a gradient boosting framework. Diagnostic models were developed, validated and reported according to the TRIPOD guidelines ^63^ (https://www.tripod-statement.org/). Gradient Boosting trees ^70, 71^, a decision-tree-based ensemble model, differ fundamentally from conventional statistical techniques that aim to fit a single model using the entire dataset. Such ensemble approach improves performance by combining strengths of models that learn the data by recursive binary splits, such as trees, and of “boosting”, an adaptive method for combining several simple (base) models. At each iteration of the gradient boosting algorithm, a subsample of the training data is selected at random (without replacement) from the entire training data set, and then a simple base learner is fitted on each subsample. The final boosted trees model is an additive tree model, constructed by sequentially fitting such base learners on different subsamples. This procedure incorporates randomization, which is known to substantially improve the predictor accuracy and also increase robustness. Additionally, boosted trees can fit complex nonlinear relationships, and automatically handle interaction effects between predictors as addition to other advantages of tree-based methods, such as handling features of different types and accommodating missing data. Hence, in many cases their predictive performance is superior to most traditional modelling methods. Additional gain of these algorithms is the various loss functions that can be applied. Using the softmax loss function, we explicitly estimated the class conditional probabilities, which allow us to demonstrate the performance of each of the classifiers both as “soft-classifiers” (i.e., predicting class probabilities) and “hard-classifiers” (i.e., setting a probability threshold and predicting a class). The former approximates a continuous number as output - the class conditional probabilities - and then performs classification based on these estimated probabilities. In contrast, hard classifiers output a discrete number as the decision - directly targeting the classification decision boundary, without producing the probability estimation.

A gradient boosting classifier was developed with a feature set of 132 miRNA predictors. Dataset was partitioned to training-set (75%) and validation-set (25%) which was used as held-out data. The training-set was cross-validated during training with stratified 3-fold cross validation. An ROC was generated for each of the folds and individual and mean AUCs were calculated along with 95% confidence intervals.

The chosen hyper-parameters: ccp_alpha=0.0, learning_rate=0.5, max_depth=8, max_features=0.45, min_samples_leaf=14, min_samples_split=8, n_estimators=100, subsample=0.45 and tol=0.0001.

### miRNA Predictor Selection by Recursive Feature Elimination (RFE)

For selecting the most predictive features during prediction model development, we used *Recursive Feature Elimination* (RFE) algorithm, an efficient recursive approach for eliminating features from a training dataset with 3-fold cross validation. ExtraTreesClassifier algorithm is used in the cross-validated RFE procedure, with hyper-parameters: criterion="entropy", max_features=0.9, and n_estimators=20. RFE works by iteratively removing features and using model accuracy to identify which features contribute the most to prediction. Tree-based importance scores of 132 miRNAs were used in order to rank features, and thus reduced the dimension of miRNA measurements needed for prediction by ∼90% (13 miRNA features in a model in total). Additionally, the final model included age and sex as predictors (resulting in a of total 15 predictors).

### Feature Importance and SHAP Analysis

Although gradient boosting tree models are complex models, they can automatically provide an approximation of feature importance from the trained boosted trees. A miRNA predictor is assigned with an importance score in every single tree, where the Gini purity index is used to assess split points in the tree. The score of a feature is calculated based on the amount of improvement in the Gini index achieved by split points that include the feature, weighted by the number of observations in that node. The final importance score of a feature is calculated by an average across all decision trees within the final model.

For local interpretability of the predictive model, we used SHapley Additive exPlanations (SHAP) ^72^, the current state of the art in Machine Learning explainability tools. SHAP provides estimates and visualizations to infer what decisions the model is making. This is achieved by quantifying the contribution that each feature brings to each prediction made by the model.

### Linear regression model

A linear classifier (Logistic Regression) was developed with a feature set of 95 miRNA predictors, that were differentially expressed (adjusted p-value ≤ 0.05) between FTD cases and controls. Each miRNA was binarized by its mean value. Training and validation were identical as in the gradient boosting classifier: Dataset was partitioned to training-set (75%) and validation-set (25%) which was used as held-out data. The training-set was cross-validated during training with stratified 3-fold cross validation. An ROC was generated for each of the folds and individual and mean AUCs were calculated along with 95% confidence intervals. The chosen hyperparams for Logistic Regression: L2 penalty, tol=1e-4, fit_intercept=True, solver=’lbfgs’, max_iter=500.

## Results

We sought to determine the overall diagnostic capability of miRNA measurements in FTD. To this end, we based our study on analysis of plasma miRNA expression and the development of computational diagnostic models. The cohort included a total of 293 participants, enrolled between 2009 and 2018. Summary of participants’ basic characteristics is shown in Table 1. Since the non-neurodegenerative controls were collected in two centers, we first verified that they could have been sampled from a single population and can therefore be considered a single cohort. We estimated the Kullback–Leibler divergence, a measure of the difference between two probability distributions. This analysis indicated only a small difference between the two sets in question (KL = 0.293, Figure S1), that is further visualized by t-distributed stochastic neighbor embedding (t-SNE) analysis of the cohorts.

### Differential expression of miRNAs

Out of the >2000 miRNA species that were aligned to the human genome, only 132 fulfilled QC criteria of an average UMI count ≥ 100 across all samples and non-zero counts in all samples (see Methods). Next, we quantified the differential miRNAs that may distinguish FTD from controls. Ninety-five miRNAs were differentially expressed between plasma of patients with FTD and controls, with an adjusted p-value<0.05 (Figure 1A). Additional analysis of only subsets of C9ORF72-FTD cases (Figure 1B), FTD females (Figure 1C) or FTD males (Figure 1D) vs. relevant controls revealed that the miRNA signature was comparable between the full FTD cohort and subcategories (Table S2: Source Data Fig. 1). This was also true for subsets of FTD patients with predicted TDP or Tau pathology (Figure S2).

**Figure 1.**
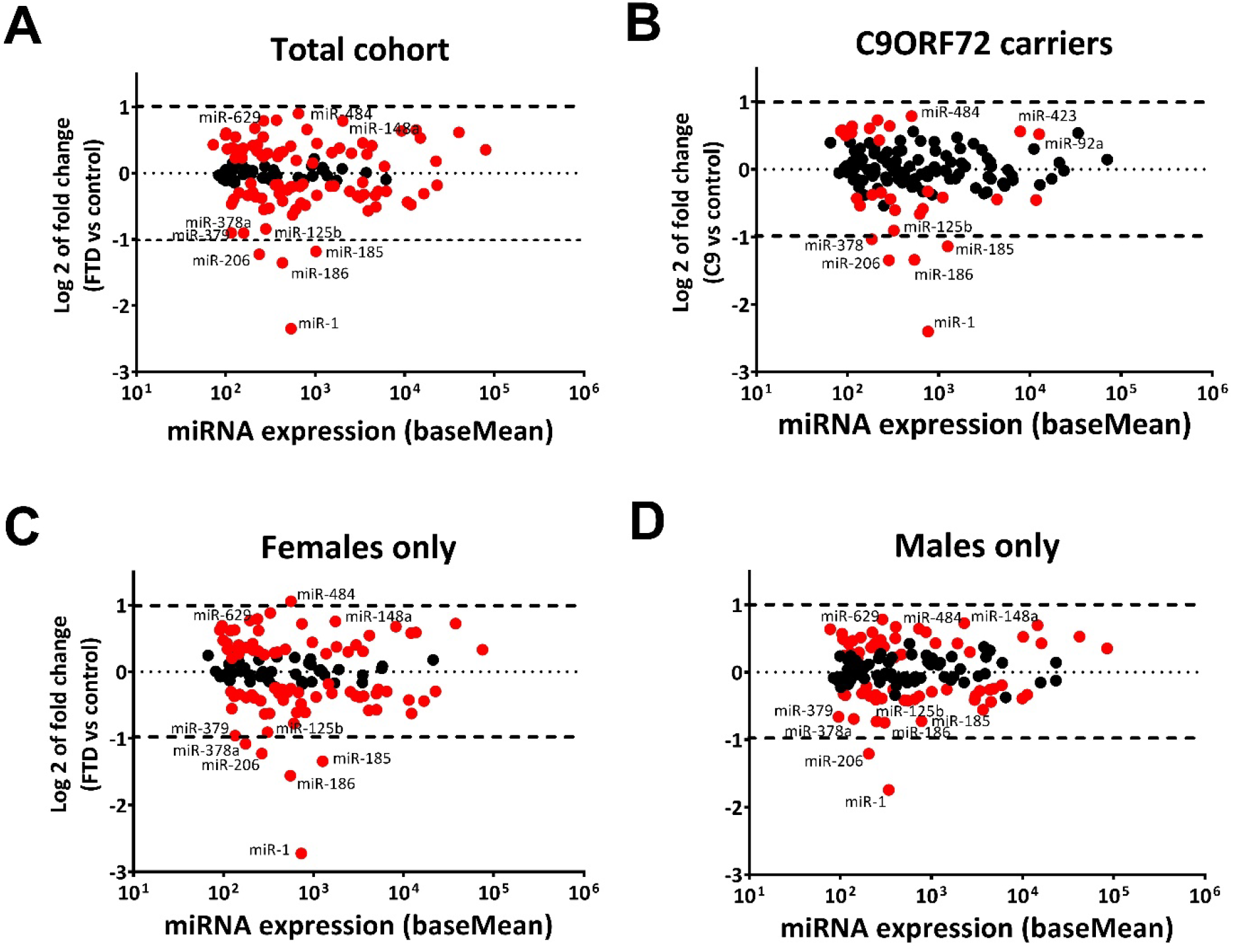
miRNA signature associated with FTD. MA plot of differential miRNA expression in plasma of patients with FTD vs non-neurodegenerative healthy controls in the **(A)** total cohort of FTD patients and controls (N=168 and N=125, respectively). **(B)** C9ORF72 mutations carriers only (N=18) vs healthy controls (N=125). **(C)** Female patients with FTD and healthy female controls (N=58 and N=85, respectively) **(D)** male patients with FTD and healthy male controls (N=110 and N=40, respectively). Log 2 transformed fold change (y-axis), against mean miRNA abundance (x-axis). Red: significantly changed miRNAs (adjusted p<0.05, Wald test). Black: miRNAs showing insignificant change.

### Development of machine learning classifier for the diagnosis of FTD

We established a diagnostic prediction model for FTD on a randomly selected training set of 135 FTD cases and 84 controls, comprising 75% of the total cohort (168 cases, 125 controls). For model validation, the remaining 25% of the data were held out as a replication cohort (33 FTD, 41 control samples).

The 132 miRNAs were tested as potential predictors, using an ensemble machine learning approach for ranking miRNAs predictive value in the diagnosis of FTD vs. individuals that did not suffer from neurodegeneration and were considered healthy. For selecting the most predictive features during model development, we used Recursive Feature Elimination (RFE), an efficient multivariate approach that iteratively removes miRNAs (features) and identifies those that contribute the most to prediction accuracy. Cross validated RFE on the training set with Extra Tree classifiers, obtained a set of 13 miRNAs with highest feature importance (Figure 2A). Age and sex were added as predictors in the model, in addition to the selected 13 miRNAs features.

**Figure 2:**
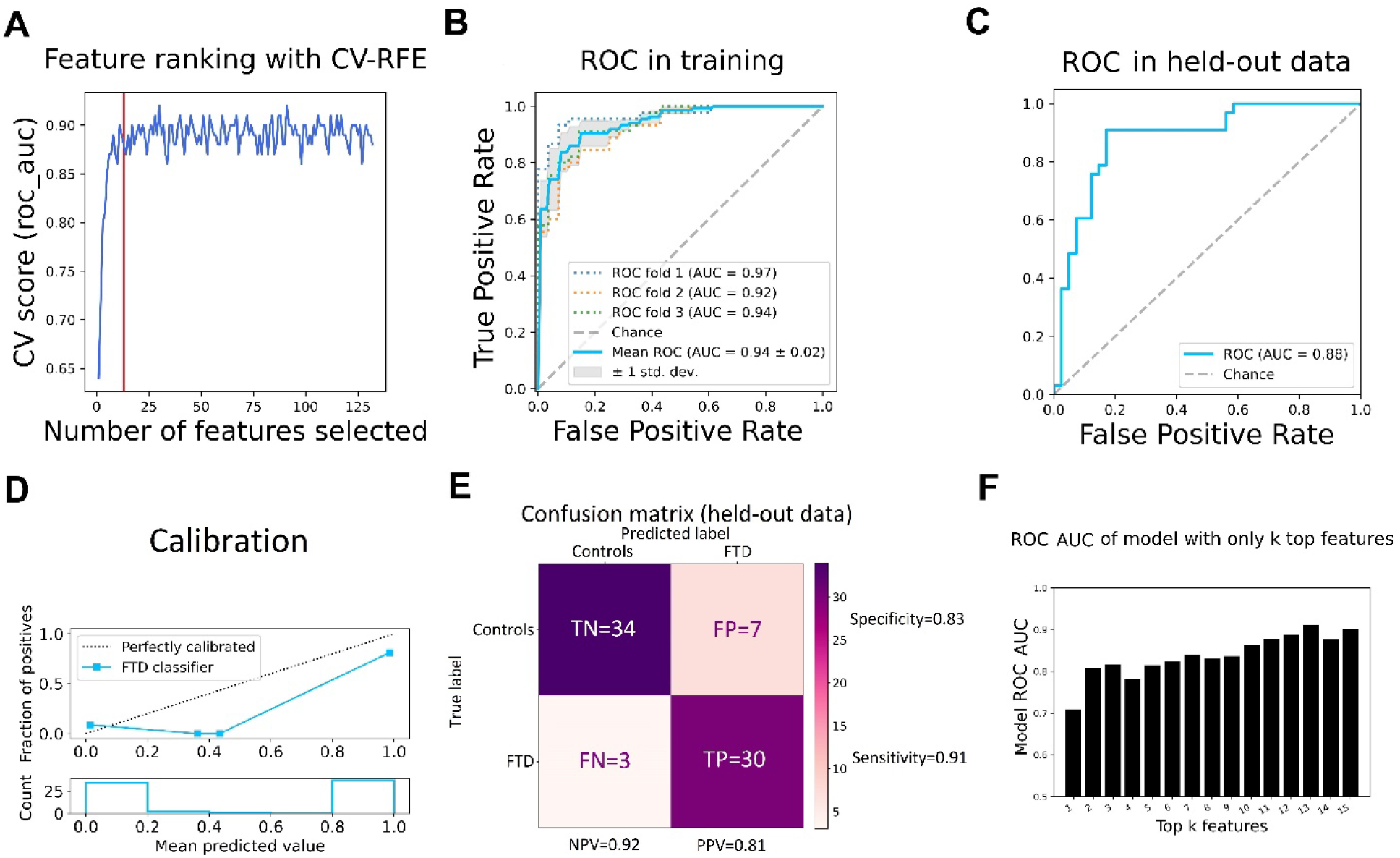
Diagnosis of FTD by a distinctive cell free miRNA signature. **(A)** Mean accuracy under 3-fold cross validation (AUC ROC, y-axis) as a function of the number of plasma miRNAs used in the FTD prediction model. Increasing numbers of miRNA features (x-axis) successively selected in the recursive feature elimination process reveals a saturation at 13 miRNA features (red line). Final model included 15 features (including age and sex as covariates). **(B)** ROC curves in the training set: true positive rate (y-axis) vs. false positive rate (x-axis). Mean values and variance of data from 219 samples with 3-fold cross validation. Mean AUC 0.94 ± 0.02. 95% CI is 0.92-0.96. **(C)** Performance and generalizability on replication held-out data revealed by an ROC curve with AUC of 0.88. **(D)** Reliability diagram, plotting truly observed fraction of cases (upper plot, y-axis) vs. predicted probability by the prediction model (upper plot, x-axis), in five probability bins, reveals a sufficiently calibrated model on the held-out set. Lower plot shows the number of individuals at each of the five predicted probability bins. **(E)** Confusion matrix showing the prediction errors on held out dataset. At a probability threshold of 0.6, we observed 0.92 Negative predictive value (NPV), 0.81 Positive Predictive Value (PPV/precision), 0.91 recall (sensitivity) and 0.83 specificity. In addition, a 0.17 False Positive Rate is obtained over 0.08 False Omission Rate and 0.19 false discovery rate. **(F)** AUC ROC of a model trained with only a subset of top 15 most predictive features, reveals a stable performance with the selected final features.

The prediction model presented a mean receiver operating characteristic area under the curve (ROC AUC) of 0.94 under internal cross validation on training dataset (Figure 2B). Furthermore, the 13-miRNA-based predictor was capable of predicting the diagnosis of FTD in an independent held-out (replication) cohort of 74 individuals with an ROC AUC of 0.88 (Figure 2C). Additional analysis reveals that the model is well calibrated and satisfactorily performs on held-out data (Figure 2D). At a defined probability threshold (0.6), the classifier exhibits a 0.81 precision (PPV) over 0.92 negative predictive value (NPV) with a 0.91 recall (sensitivity), a 0.83 specificity and a 0.08 false omission rate over 0.19 false discovery rate values (Figure 2E). Furthermore, a family of models trained with only a subset of the 15 most predictive features, displays a stable AUC ROC performance and reassures that the selected final features are suitable (Figure 2F). In summary, we determined 13 miRNAs that are able to call the diagnosis of FTD with high accuracy. We have further compared the non-linear machine-learning strategy (with 13 miRNA features selected by multivariate RFE, Figure 2) to a logistic regression model with univariate feature selction (Figure S3). A subset of 95 miRNAs out of the 132 passing QC, were differentially expressed between FTD and controls (passing a threshold of p<0.05 after correction for multiple hypotheses) and therefore were used in the logostic regression as features. Similarly, age and sex were also included as predictors. The linear model with 95 differentially-expressed miRNAs, age and sex was inferior to the gradient boosting classifier model, particularly by sensitivity (0.73 vs 0.91) and negative predictive value (0.8 vs 0.92) (Figure S3). Moreover, the non-linear approach showed better robustness in the different training folds and also outperforms the linear model in utilizing a significantly smaller number of miRNA features (15 vs. 97), to obtain its accuracy.

We then sought to better understand the machine-learning-based non-linear prediction model by investigating the relative effect of each individual miRNA. Therefore, we utilized post-hoc SHapley Additive exPlanations (SHAP) feature importance analysis to uncover the contribution of individual miRNAs to the FTD diagnostic predictor (Figure 3A, B). Extreme SHAP values inform that the model predicts a more likely FTD (positive values) or healthy control (negative values). The key predictors revealed by SHAP, which contribute the most to calling FTD vs controls, are multiple sclerosis-associated miR-629 ^73, 74^, brain enriched miR-125b ^75^ and the astrocyte-derived exosomal miR-361 ^76^. We further tested each individual miRNA as a single predictor of FTD diagnosis. A model with only miR-423-5p or miR-125b-5p presented the highest AUC values of ∼0.69 (Table 2). Boxplots depicting the underlying distribution of each of the 13 miRNA predictors, along with scatter plots of the expression values of individual subjects in these miRNAs, confirm the non-linear relationship between miRNA expression and the phenotype, for some miRNAs (e.g., miR-107 and miR-26a), while other miRNAs can be divided to low/high values predominated by either controls or FTD cases (e.g., miR-629-5p, Figure 4). These observations emphasize the power of a model, which is able to take into account non-linear relationship between features (miRNAs).

**Figure 3:**
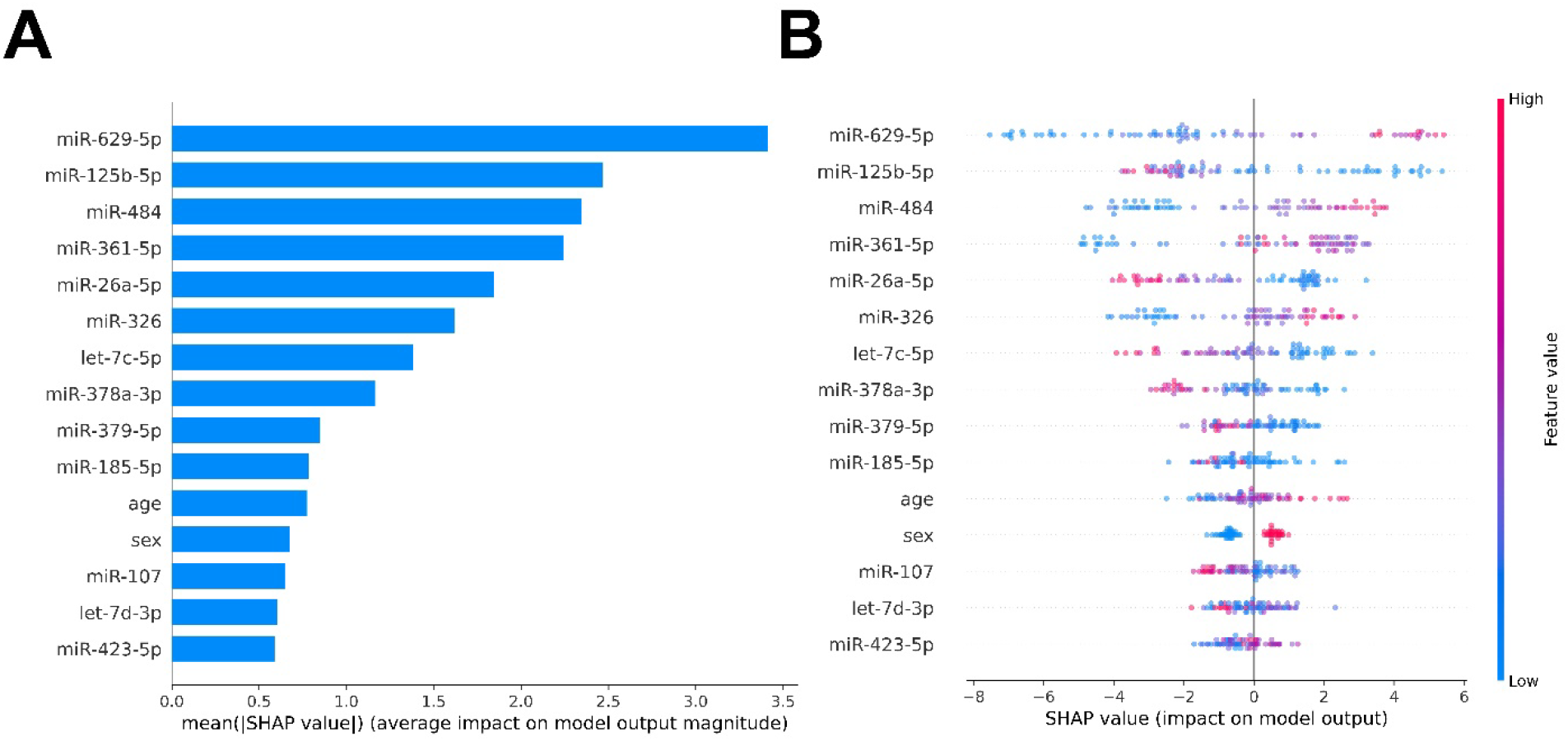
The contribution of individual miRNAs to the predictor of FTD diagnosis. **(A)** Mean absolute SHapley Additive exPlanations (SHAP) values break down the impact of specific miRNAs on FTD disease non-linear predictor output in the held-out cohort. **(B)** Illustration of the relationship between the miRNA levels (low – blue to high – red), SHAP values and the impact on the prediction in the held-out cohort. Positive or negative SHAP value lead the model to a more likely FTD or healthy control predictions, respectively.

**Figure 4.**
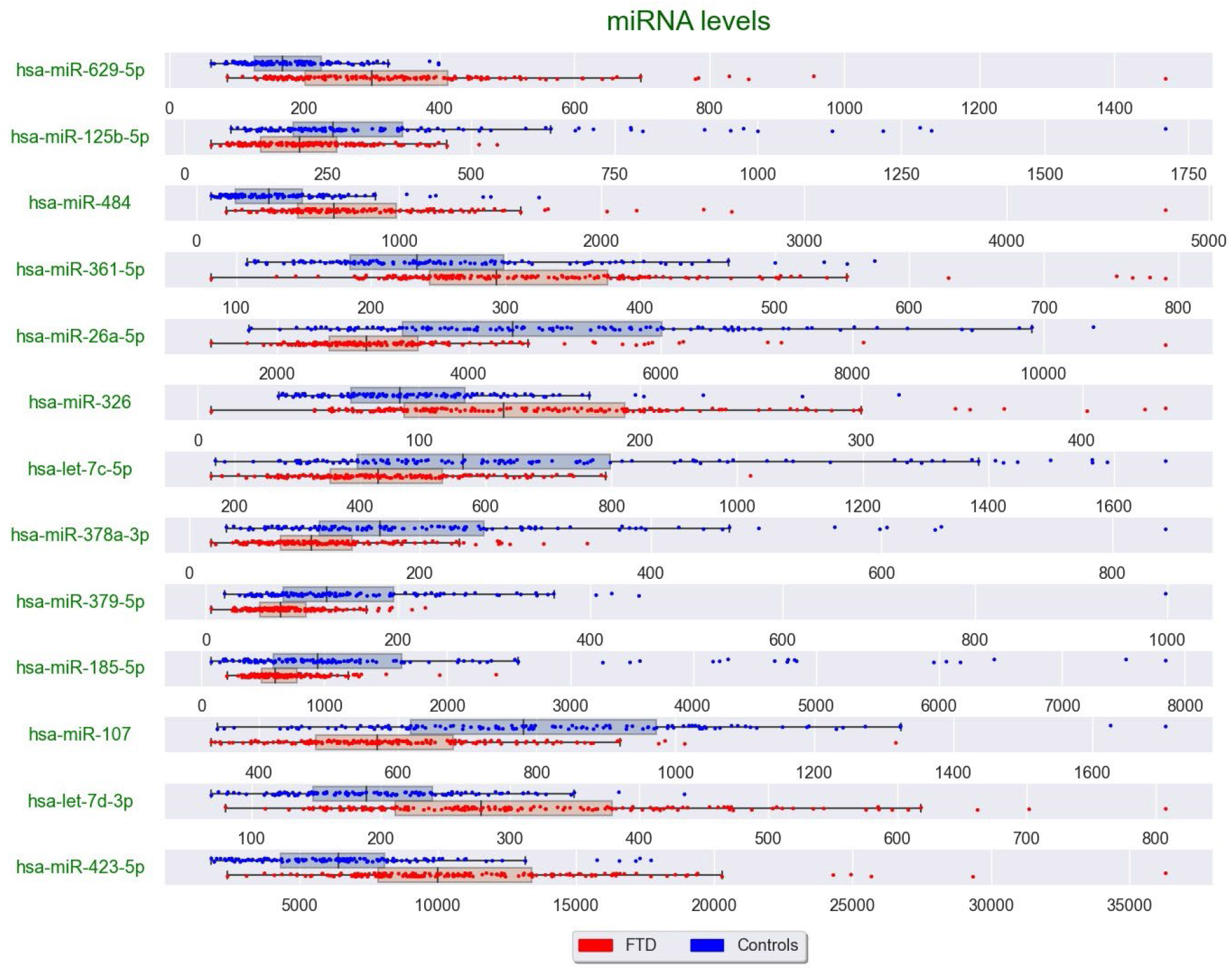
Distribution of the top 13 miRNA predictors in patients with FTD and controls. Box plots with complement <u>scatter</u> plots of the 13 miRNA features predicting FTD, depicting expression values of all cohort along with distribution of each miRNA. Blue dots represent the levels of a given miRNA measured in healthy control participants. Red dots represent the levels of a given miRNA measured in patients with FTD. Box – two central data quartiles, with a line at the median (Q2). Whiskers extend to show the rest of the distribution, except for points that are determined to be outliers using a method that is a function of the inter-quartile range.

**Table 2.** Predictive power of selected miRNA features, when used as a single predictor for FTD on held-out data. Among the most predictive features are miRNAs expressed in the brain, such as miR-26a-5p, miR-125b-5p and let-7c-5p.

| Predictor | Single Feature AUC |
| --- | --- |
| miR-423-5p | 0.69 |
| miR-125b-5p | 0.67 |
| miR-26a-5p | 0.66 |
| sex | 0.66 |
| miR-326 | 0.65 |
| miR-185-5p | 0.64 |
| age | 0.64 |
| miR-629-5p | 0.63 |
| miR-484 | 0.62 |
| let-7d-3p | 0.62 |
| miR-107 | 0.62 |
| let-7c-5p | 0.61 |
| miR-361-5p | 0.59 |
| miR-379-5p | 0.56 |
| miR-378a-5p | 0.54 |

Significant pairwise differences were noted in miR-107 levels in both bvFTD and SD vs other FTD subtypes (primary progressive aphasia, corticobasal syndrome, progressive supranuclear palsy and FTD with motor neuron disease. ANOVA: p=0.009, bvFTD vs other FTD subtypes: p=0.01, SD vs other FTD subtypes: p=0.02, Figure S4C). In addition, significant pairwise differences in miR-26a levels were found in both bvFTD (p=0.04) and PNFA (p=0.02) vs other FTD subtypes (ANOVA: p=0.024, bvFTD vs other FTD subtypes: p=0.04, PNFA vs other FTD subtypes: p=0.02, Figure S4F).

None of the other miRNAs showed differences between clinical subtypes (Figure S4), between different mutation carriers, or between mutations carriers and patients with no known FTD mutations (Figure S5). In conclusion, a non-linear signature based on only 13 miRNAs is able to call the diagnosis of FTD with high accuracy and generalizability, for all FTD subtypes and independent of the underlying genetic background.

Finally, we tested the performance of plasma miRNA classifiers that were reported in previous works ^56, 58^, relative to the classifier we have reported here. We replicated the logistic regression model (with L2 regularization^77, 78^) from the study of Kmetzsch et al. ^58^ on our data. Taking miR-34a-5p, miR-345-5p, miR-200c-3p and miR-10a-3p as features, displayed inferiority to our model. Moreover, these miRNAs were not differentially expressed between cases and controls in our study. Next, we replicated the "microRNA pair" approach from the study of Sheinerman et al. ^56^: miR-335/let-7e, miR-99b/let-7e and miR-9-3p/miR-181a and assessed the ROC curves, showing they are inferior to our non-linear classifier in distinguishing between FTD and controls (Figure 5). Thus, when tested in comparison to reported miRNA classifiers from the literature, our panels of miRNA and non-linear classifier perform better, with AUC values of 0.9.

**Figure 5.**
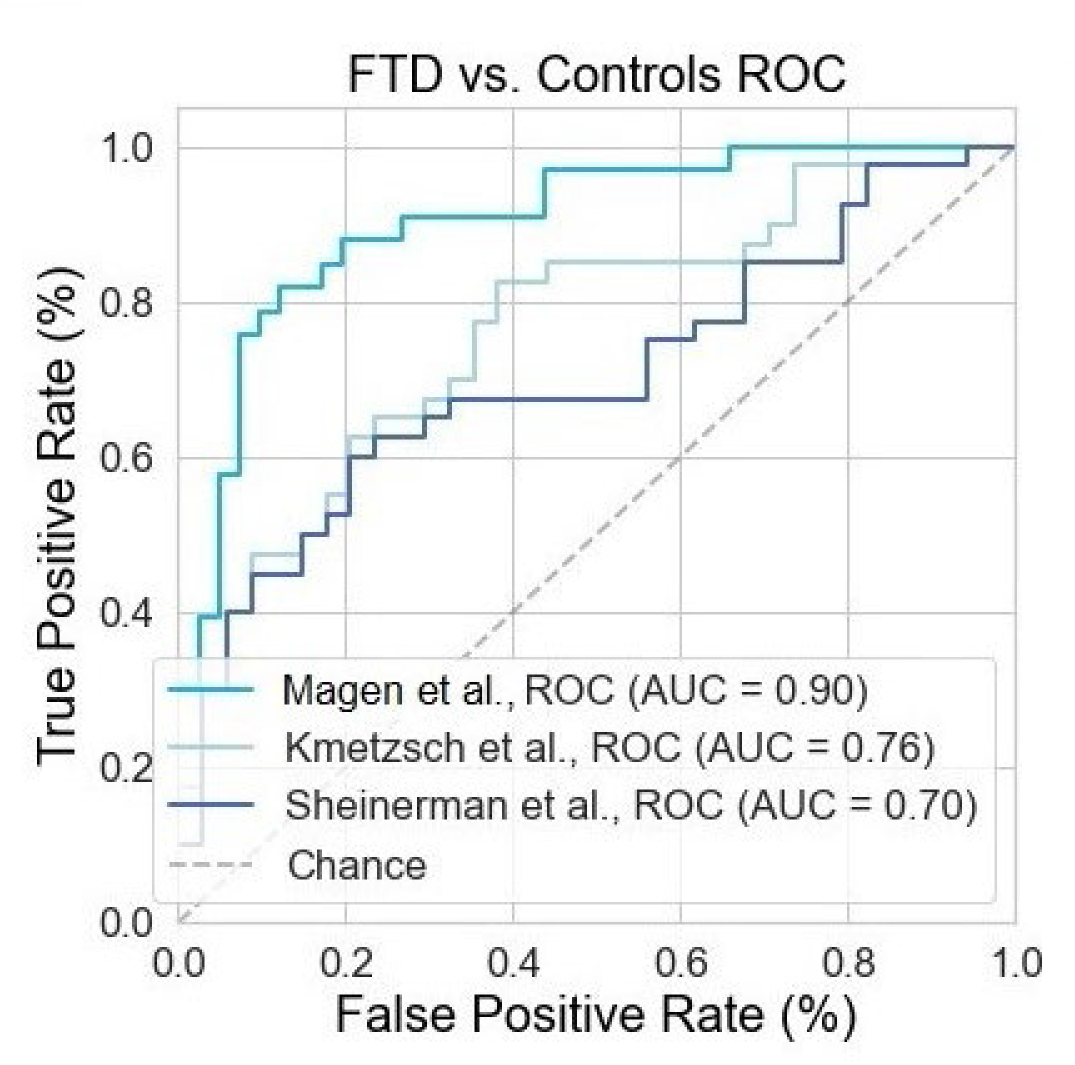
AUC values for different miRNA classifiers for discriminating FTD vs controls in the held-out data. The classifiers were either the 13 selected in the FTD model in our data, a combination of miR-34a-5p, miR-345-5p, miR-200c-3p and miR-10a-3p [from], or a combination of miR-335/let-7e, miR-99b/let-7e and miR-9-3p/miR-181a [from];

## Discussion

The need to facilitate the diagnosis of FTD in the face of clinical heterogeneity raises the hope for new effective biomarkers. Circulating miRNAs hold a fascinating potential as diagnostic biomarkers that were not fully explored, including for brain disorders ^50^. We recently demonstrated the power of miRNAs in prognosis and prediction of clinical response to therapy in motor neuron diseases ^51, 52^. In the present work, we sought to discover diagnostic biomarkers for FTD. By using an unbiased, next generation sequencing approach and advanced computation in a discovery cohort of 219 subjects and 74 additional subjects for a replication study, we overcame limitations of past works in developing biomarkers for FTD. A recursive non-linear approach was utilized to find the smallest set of miRNA features that obtain the highest accuracy of the prediction model, leading to a signature of 13 miRNAs only. Thus, our miRNA-based binary classifier is more accurate and robust than published miRNA-based predictors ^56, 58^.

The use of gradient boosting trees, an ensemble learning approach, allows discovering nonlinear relationships between miRNAs and disease status that gained affirmation by cross validation in the training dataset. Furthermore, our model is externally validated on held-out data, which was not used during feature selection and model development, according to the TRIPOD guidelines ^63^. SHAP analysis ^72^ further unfolds the relative contribution of individual features to the predictive model. Of note, the performance of the ensemble learning was superior to that of a logistic regression model, and enabled the use of only 15 features, instead of 97, for prediction.

According to the human miRNA tissue atlas ^75^, most of the selected miRNAs are CNS-enriched (let-7c-5p, miR-26a-5p, miR-107, miR-125b-5p), suggesting that they might be directly involved in the disease state of the CNS. miR-26a, miR-326, miR-484 and miR-361 were associated with FTD diagnosis in our data and with cognitive deficits or Alzheimer’s disease (AD) in other analyses ^79–85^. In mice, miR-326 inhibited Tau phosphorylation^80^, a hallmark of FTD as well as AD, supranuclear palsy (PSP), conticobasal syndrome (CBD) and chronic traumatic encephalopathy (CTE)^86^. Brain-enriched miR-107 was also implicated in AD ^87–92^. Changes in blood levels of miR-326, miR-26a, and miR-629 are associated with multiple sclerosis (MS), a condition characterized by demyelination ^93–100^. Moreover, serum miR-629 was negatively correlated with MS patient brain volume and lesion severity, respectively ^101^. Thus, some of the miRNA predictors proposed here are associated directly or indirectly with other degenerative brain diseases.

## Conclusion

### Strengths and Limitations

We have found specific molecular miRNA patterns that can contribute to the diagnosis of FTD. Therefore, the work encourages testing if circulating miRNAs biomarkers can establish a cost-effective screening approach to increase speed or precision in the diagnosis of suspected FTD. Early-stage diagnosis may be useful towards design of prospective clinical trials. More broadly, the findings demonstrate the importance of integrating machine learning into clinical biomarker studies, addressing nonlinearity and exposing otherwise cryptic disease-associated signals. Finally, while ongoing biomarker studies in FTD highlight protein markers such as NfL and Tau, combined protein-RNA markers may present increased accuracy, as we previously showed^52^.

We would like to put forward a few notable limitations of our study: First, patients in our cohorts were most likely recruited in different phases of the disease, which results in significant phenotypic heterogeneity. At the same time, since we had no record of disease severity, we could not stratify patients by their disease phase and depict stage-dependent changes in miRNAs. Additionally, we did not find any miRNAs, which can differentiate between FTD-Tau and FTD-TDP, which might result from small, underpowered subset cohorts or could highlight a wider role for these novel biomarkers within dementia syndromes. We hope that in the future, larger cohorts can be used to reveal such differences. Finally, a long route is expected from this initial study and until miRNA can be used in personalized diagnosis. These steps shall include prospective studies, quantitative calibration of absolute miRNA concentrations and simple bed-side methods for quantification of miRNAs.

### Unanswered questions

It remains to be determined why our findings are discrepant with conclusions of past studies ^53–60, 62^. For studies in other bodily fluids the answer might be trivially-related to the different biofluid composition^53, 57, 60^. Furthermore, we emphasize the progress we presented in our study in terms of power (larger cohort) and the unbiased analysis by next generation microRNA sequencing, that contrasts past biased choices of miRNA candidates. A second unanswered question is why defined nonlinear patterns are strongly predictive of disease states. Future works should address the differential diagnosis between FTD and other dementias, such as Alzheimer’s disease. Lastly, protein based markers, such as neurofilaments, lack specificity to particular neurodegenerative diseases. However, it is plausible that miRNAs might demonstrate a disease-specific pattern in the circulation. Future studies with large AD and FTD cohorts might address this hypothesis.

### Recommendations

In our study we took care to keep a held-out cohort for external validation. To substantiate our work and towards clinical diagnostic usage, it is warranted to validate the predictor by testing it on an independent cohort of different ethnicity and create means for quantification of absolute miRNA concentrations. In addition, miRNA levels should be compared in follow up works, and combined with other experimental circulating markers of neurodegeneration such as neurofilaments. In addition, miRNAs could be explored also as prognostic markers in FTD and in predicting disease severity.

### Availability of supporting data

The data that support the findings of this study are available from the corresponding author upon reasonable request.

## Appendix

### IDs of 34 subjects removed due to age-based QC

’CTRL_rep9’, ’CTRL_rep10’, ’CTRL_rep13’, ’CTRL_rep19’, ’CTRL_rep16’, ’CTRL_rep18’, ’CTRL_rep20’, ’FTD30’, ’CTRL_rep17’, ’CTRL12’, ’CTRL13’, ’CTRL2’, ’CTRL5’, ’CTRL56’, ’CTRL57’, ’CTRL58’, ’CTRL68’, ’CTRL100’, ’CTRL111’, ’CTRL20’, ’CTRL22’, ’CTRL23’, ’CTRL24’, ’CTRL27’, ’CTRL29’, ’CTRL33’, ’CTRL48’, ’CTRL50’, ’CTRL54’, ’CTRL60’,’CTRL62’, ’CTRL73’, ’CTRL82’, ’CTRL83’.

## Data Availability

All data produced in the present study are available upon reasonable request to the authors

## Acknowledgements

We thank Vittoria Lombardi (UCL) for technical assistance. We acknowledge patients with FTD, and healthy volunteers for their contribution and ALS biomarkers study co-workers for biobanking, which has made this study possible (REC 09/H0703/27). We also thank the North Thames Local Research Network (LCRN) for its support. EH is the Mondry Family Professorial Chair and Head of the Nella and Leon Benoziyo Center for Neurological Diseases and of the Andi and Larry Wolfe Center for Neuroimmunology and Neuromodulation.

## Authors’ contribution

**I**.M., P.F., J.D.R. and E.H. conceived research.

I.M., N.S.Y. and E.H. analyzed the data.

A.M. and J.D.R. established cohort, gained ethical approval and collected human samples for research.

Y.B., C.H. and I.S. assisted research.

I.M., N.S.Y., P.F., A.M., J.D.R. and E.H. wrote the manuscript, with comments and final approval by all other authors.

J.D.W. provided resources for research and input in research development.

A.M. and J.D.R. are corresponding authors for cohorts and clinical data. P.F. and E.H. are corresponding authors for all other facets of the work.

All authors read and approved the final manuscript.

## Supplementary figure and tables

**Supplementary Figure 1.**
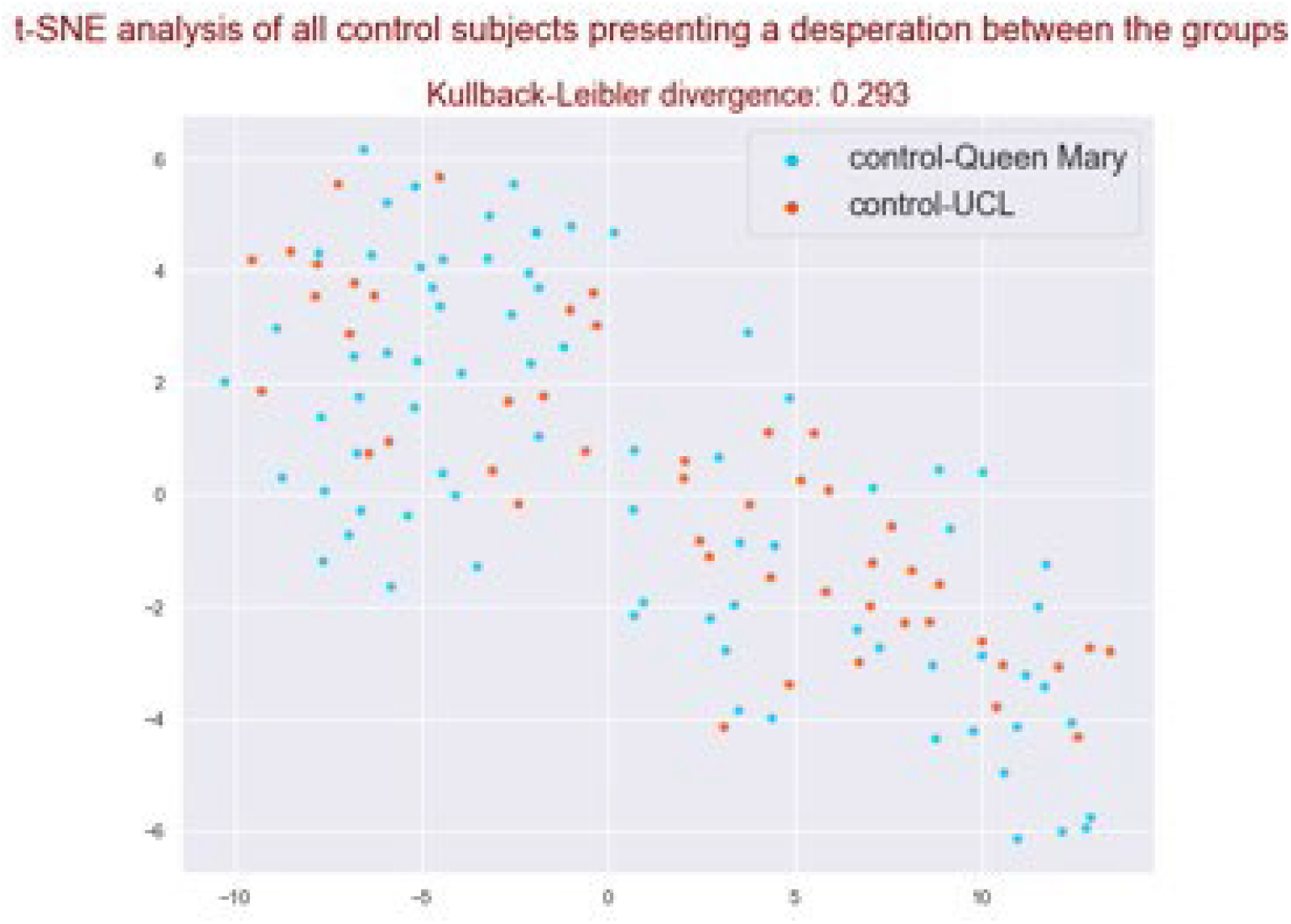
t-SNE analysis of control subjects from Queen Mary Hospital (blue) and from UCL (red). Similarity of the two control groups suggests they could have been taken from a single distribution which is justified further by an associated Kullback–Leibler divergence value of 0.293, a measure of the difference between two probability distributions.

**Supplementary Figure 2.**
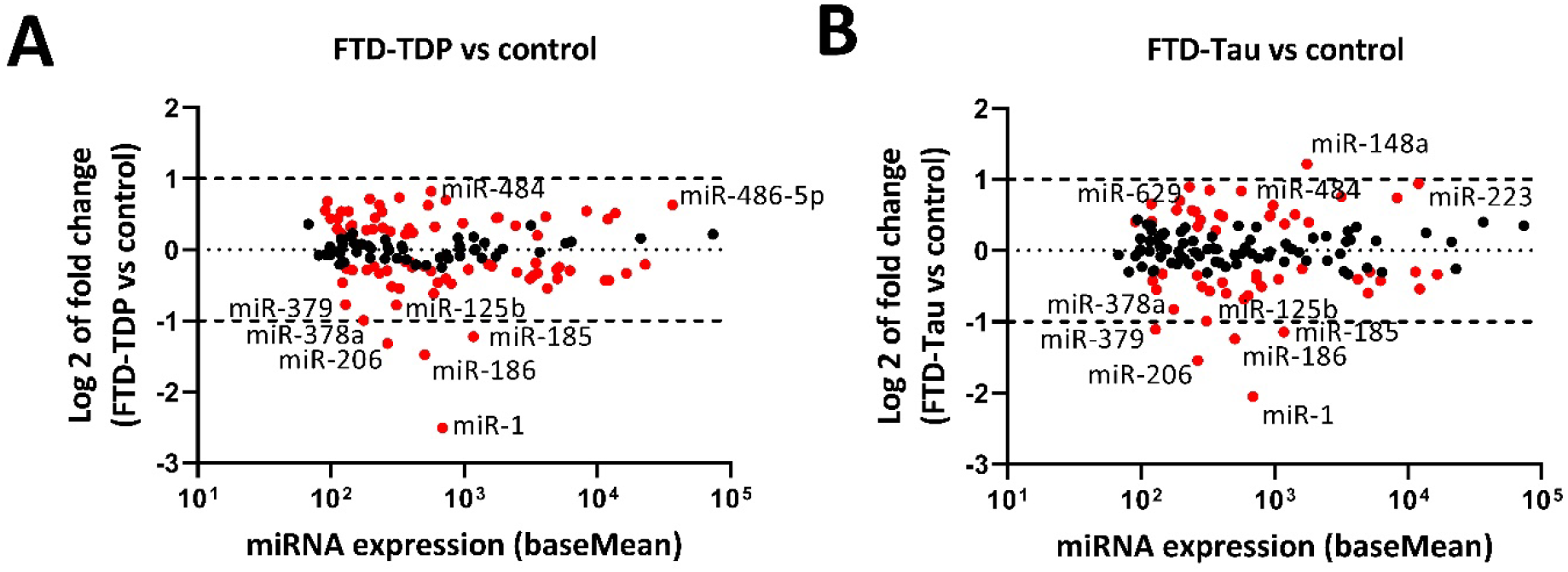
miRNA signature associated with FTD patients with predicted pathology. MA plot of differential miRNA expression in plasma of patients with predcited TDP pathology (**A; N=63**) or Tau pathology (**B; N=19**) vs non-neurodegenerative healthy controls (N=125). Log 2 transformed fold change (y-axis), against mean miRNA abundance (x-axis). Red: significantly changed miRNAs (adjusted p<0.05, Wald test). Black: miRNAs showing insignificant change.

**Supplementary Figure 3.**
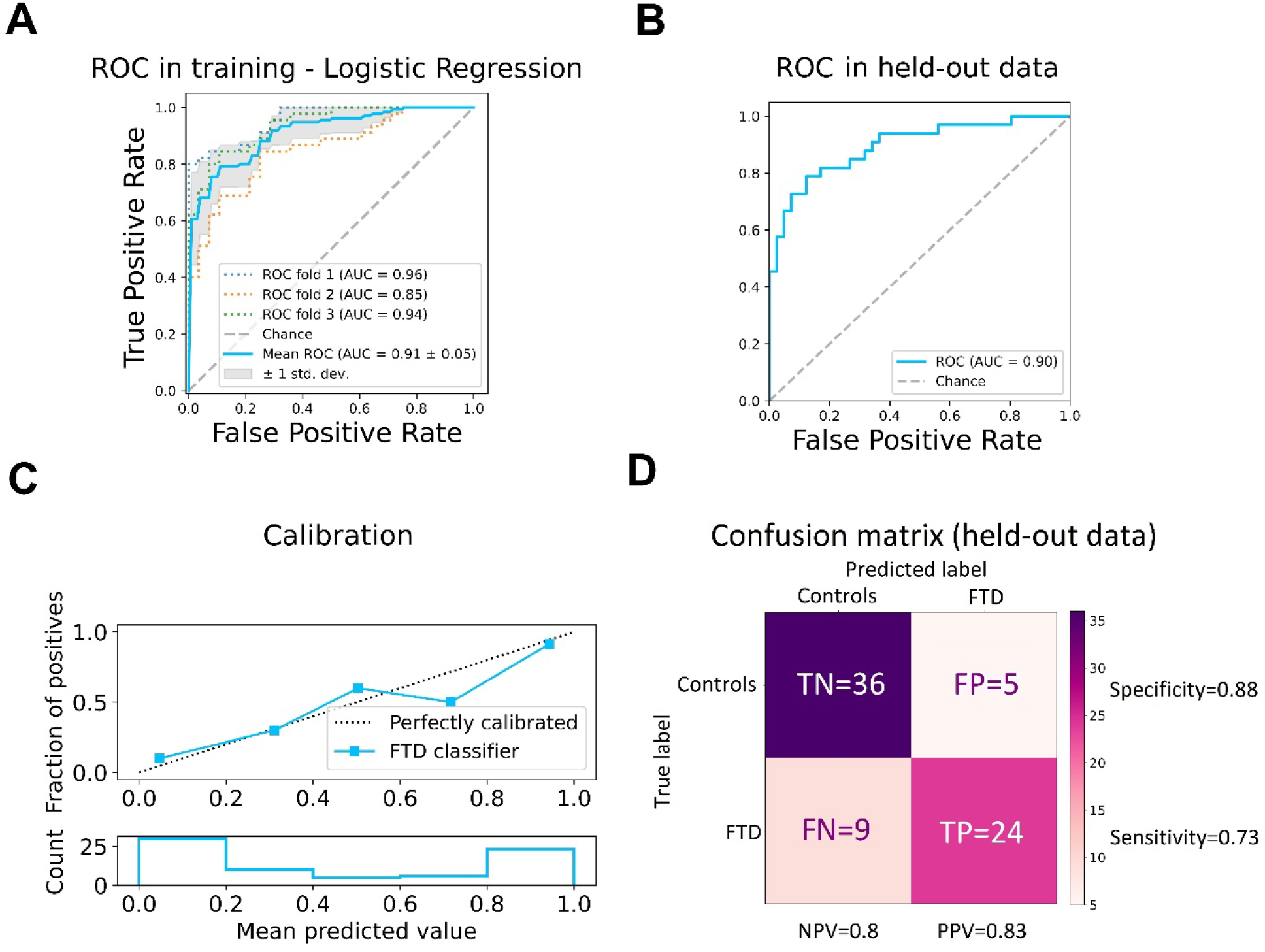
Diagnosis of FTD by age, sex and 95 differentially-expressed miRNA in a logistic regression model. ***(A)*** ROC curves in the training set: true positive rate (y-axis) vs. false positive rate (x-axis). Mean values and variance of data from 219 samples with 3-fold cross validation. Mean AUC 0.91 ± 0.05. 95% CI is 0.86-0.96. **(B)** Performance and generalizability on held-out data revealed by a ROC curve with AUC of 0.9. **(C)** Reliability diagram, plotting truly observed fraction of cases (upper plot, y-axis) vs. predicted probability by the prediction model (upper plot, x-axis), in five probability bins, reveals a sufficiently calibrated model on the held-out set. Lower plot shows the number of individuals at each of the five predicted probability bins. **(D)** Confusion matrix showing the prediction errors on held out dataset. At a probability threshold of 0.6, we observed 0.8 Negative predictive value (NPV), 0.83 Positive Predictive Value (PPV/precision), 0.73 recall (sensitivity) and 0.88 specificity. In addition, 0.12 False Positive Rate is obtained, over 0.2 False Omission Rate and 0.17 false discovery rate.

**Supplementary Figure 4.**
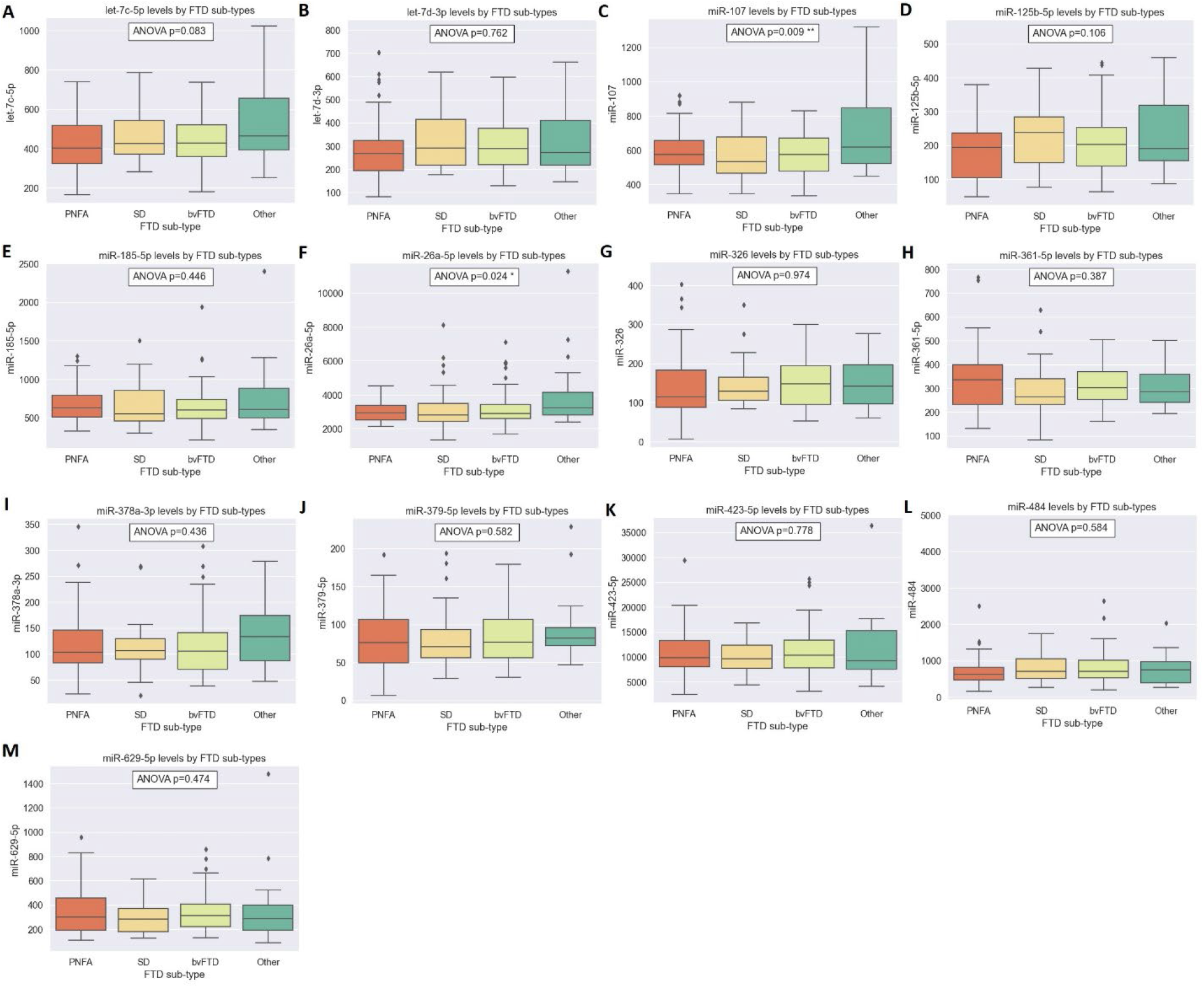
Levels of 13 miRNA predictors in different FTD subtypes. miRNA levels for PNFA (progressive nonfluent aphasia), SD (semantic dementia), behavioral FTD (bvFTD) and the following subtypes: primary progressive aphasia, corticobasal syndrome, progressive supranuclear palsy and FTD with motor neuron disease, commonly referred to as “Other”. Box plot with two central data quartiles, with a line at the median (Q2). Whiskers extend to show the rest of the distribution, except for points that are determined to be outliers using a method that is a function of the inter-quartile range. Data were analyzed by one-way ANOVA. Post-hoc Bonferroni was conducted only for miR-107 **(C)** and miR-26a-5p **(F)** which showed significant differences in ANOVA. Significant pairwise differences in miR-107 levels were found between other FTD subtypes and both bvFTD (p=0.01) and SD (p=0.02). Significant pairwise differences in miR-26a were found between other FTD subtypes and both bvFTD (p=0.04) and PNFA (p=0.02).

**Supplementary Figure 5.**
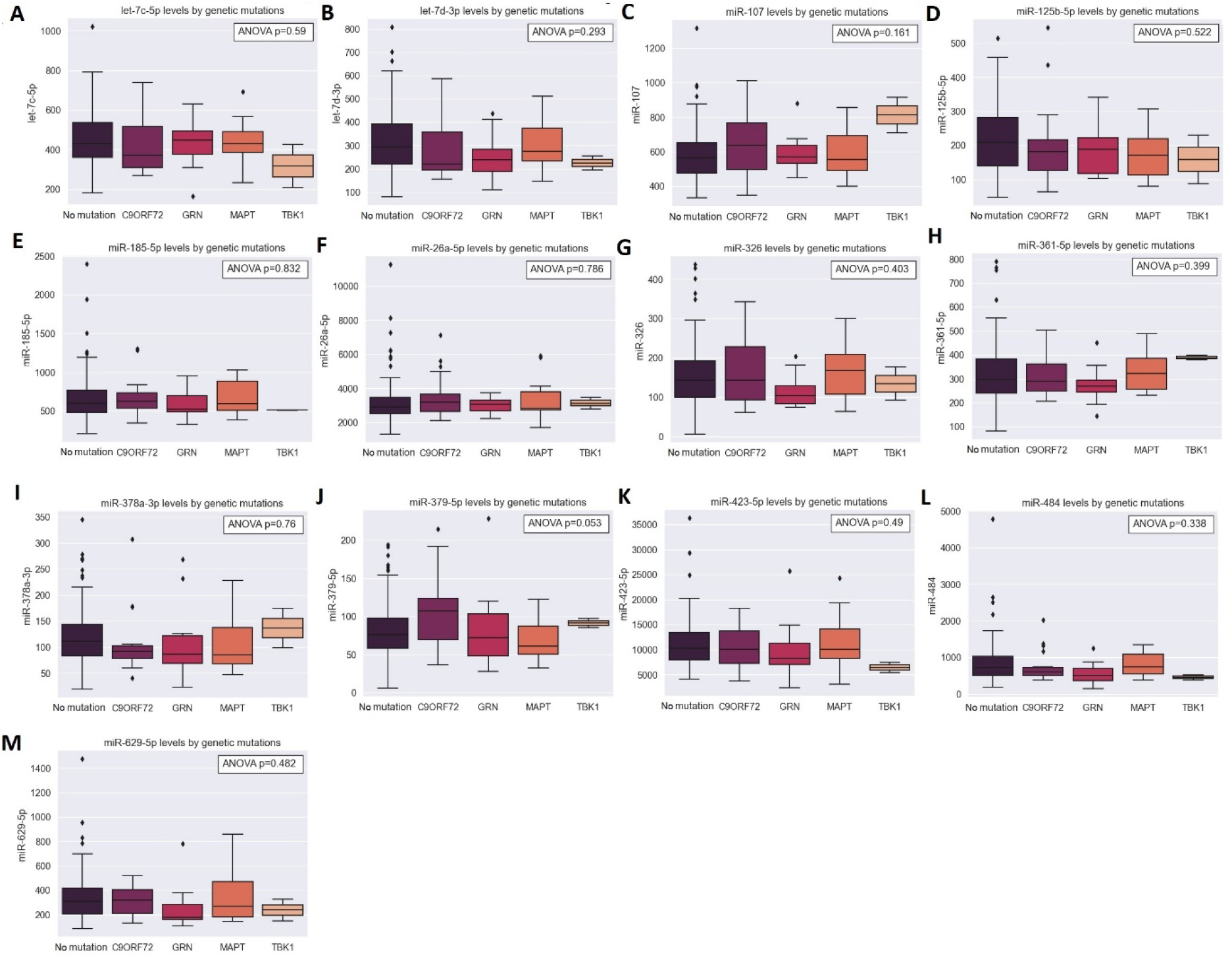
Levels of 13 miRNA predictors in mutations carriers. miRNA levels for FTD patients without known FTD mutations, or with mutations in C9ORF72, Progranulin (GRN), Tau (MAPT) and TBK1. Box plot with two central data quartiles, with a line at the median (Q2). Whiskers extend to show the rest of the distribution, except for points that are determined to be outliers using a method that is a function of the inter-quartile range. Data were analyzed by one-way ANOVA.

**Supplementary Table 1.**
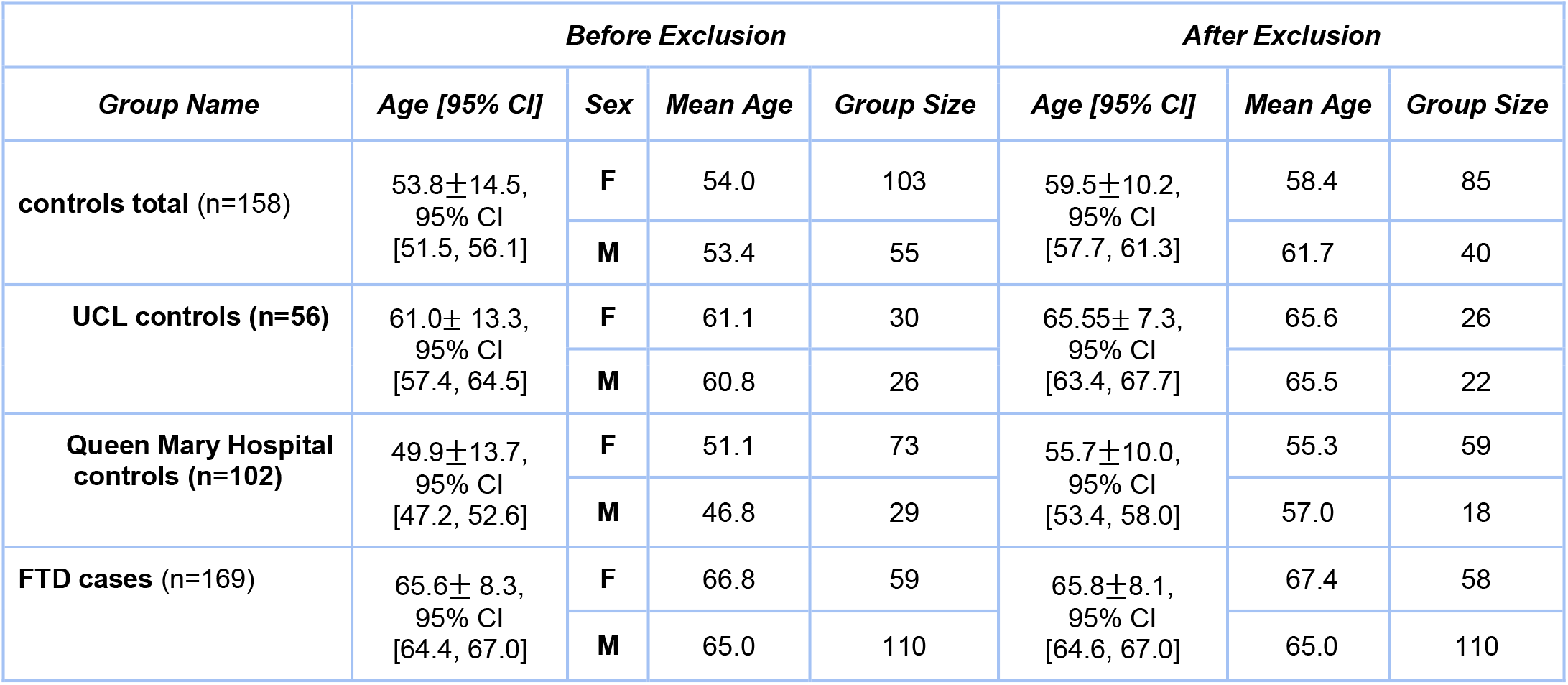
Age and sex characteristics of the cohort and exclusion by age of 34 individuals that were below 40 years of age during blood collection.

|  | *Before Exclusion* |  |  |  | *After Exclusion* |  |  |
| --- | --- | --- | --- | --- | --- | --- | --- |
| *Group Name* | *Age [95% CI]* | *Sex* | *Mean Age* | *Group Size* | *Age [95% CI]* | *Mean Age* | *Group Size* |
| **controls total** (n=158) | 53.8±14.5,<br>95% CI<br>[51.5, 56.1] | **F** | 54.0 | 103 | 59.5±10.2,<br>95% CI<br>[57.7, 61.3] | 58.4 | 85 |
|  |  | **M** | 53.4 | 55 |  | 61.7 | 40 |
| **UCL controls** (n=56) | 61.0± 13.3,<br>95% CI<br>[57.4, 64.5] | **F** | 61.1 | 30 | 65.55± 7.3,<br>95% CI<br>[63.4, 67.7] | 65.6 | 26 |
|  |  | **M** | 60.8 | 26 |  | 65.5 | 22 |
| **Queen Mary Hospital**<br>**controls** (n=102) | 49.9±13.7,<br>95% CI<br>[47.2, 52.6] | **F** | 51.1 | 73 | 55.7±10.0,<br>95% CI<br>[53.4, 58.0] | 55.3 | 59 |
|  |  | **M** | 46.8 | 29 |  | 57.0 | 18 |
| **FTD cases** (n=169) | 65.6± 8.3,<br>95% CI<br>[64.4, 67.0] | **F** | 66.8 | 59 | 65.8±8.1,<br>95% CI<br>[64.6, 67.0] | 67.4 | 58 |
|  |  | **M** | 65.0 | 110 |  | 65.0 | 110 |

